# Socioeconomic Factors and Human Knowledge and Behaviours Associated with *Aedes aegypti* abundance in Ouagadougou, Burkina Faso

**DOI:** 10.1101/2024.08.17.24310836

**Authors:** Wendegoudi Mathias Ouédraogo, Aboubacar Sombié, Federica Guglielmo, Mamadou Sermé, Hyacinthe Toé, Luca Facchinelli, Fernand Bationo, Léa Paré, Hamed S. Ouedraogo, Mafalda Viana, Antoine Sanon, David Weetman, Philip J. McCall, Athanase Badolo

## Abstract

**Background:** Since 2013, Burkina Faso has experienced regular dengue outbreaks, especially in Ouagadougou, the capital city, and Bobo-Dioulasso. Insufficient knowledge of the disease and some daily practices and attitudes of the human population may contribute to the proliferation of the vector and subsequently to dengue transmission. The objective of this study was to assess the levels of population knowledge of dengue and its primary vector, *Aedes aegypti,* and to elucidate the behavioural and environmental factors associated with the proliferation of this species.

**Methods:** A cross-sectional Knowledge, Attitude, and Practice (KAP) survey was conducted among 514 individuals, in 514 houses concurrently with a survey on *Ae. aegypti* larval sites, between September and October 2021 in three health districts (HDs) of Ouagadougou. Compound characteristics, including building materials, annual income, presence of waste, and water storage practices, were also documented.

**Results:** A proportion of94.36% of the respondents demonstrated knowledge of dengue, with 50% recognizing primary symptoms like fever and headaches, and 70.52% knowing about its transmission mode. Only 53.51% could identify *Aedes* larval containers, while 14.04% were familiar with mosquito immature stages. Certain practices were linked to increased dengue vector proliferation or biting, including water storage, open water containers, water-holding waste, and the absence of mosquito screens. The study noted a high number and diversity of potential *Aedes* larval sites with elevated Stegomyia indices (Container Index >47%, House Index >86%, and Breteau Index >230) across all HDs. Moreover, compound infestation by immature mosquitoes was correlated with tap water presence, resident count, and construction type.

**Conclusion:** Despite the dengue outbreaks in the city of Ouagadougou over the past decade and the resulting awareness campaigns, population knowledge of dengue and its vector remains limited. Therefore, there is a critical need to enhance awareness activities through campaigns aimed at changing practices and attitudes, and improving the population’s knowledge regarding the disease and its vector.

## Introduction

Dengue fever, primarily transmitted by *Aedes aegypti* and *Aedes albopictus* mosquitoes in tropical regions, is considered the most significant arboviral disease. Despite being underreported, the incidence of reported dengue cases has increased tenfold from 500,000 cases in 2000 to 5.2 million in 2019 over the past two decades, with the disease spreading to 129 countries worldwide during the same period [1]. Currently, several African countries, including Burkina Faso, are considered endemic for dengue [2], with the rapid expansion of dengue since the 1980s attributed to urbanization, globalization, and ineffective vector control [3–5].

West Africa has been identified as a potential dengue transmission hotspot [6], yet few West African nations are adequately resourced or prepared for outbreaks [7]. Burkina Faso experienced its first documented dengue outbreak in 1925, but subsequent underreporting occurred until significant contemporary outbreaks in 2016, resulting in 2,600 cases and 21 deaths [8]. In 2023, Burkina Faso witnessed one of its deadliest outbreaks ever, with 154,867 cases (70,433 confirmed) and 709 deaths (a 1% lethality rate) [9,10]. However, dengue epidemiology remains poorly investigated and documented in Burkina Faso and the West African region generally [7].

Most studies conducted in Ouagadougou on dengue vector breeding sites have consistently observed abundant and varied artificial larval containers resulting from human activities [11–13]. The typology of these breeding containers varies according to the level of urbanization, with tires and small containers being prevalent in urban areas where piped water is available but intermittent, while drums and barrels are commonly used for water storage in rural and peri-urban settings [11,12].

Household water management practices and other sociodemographic factors may serve as predictors associated with the risk of dengue vector proliferation [14,15]. Furthermore, several studies have highlighted respondents’ poor knowledge of vector breeding sites and mosquito immature stages’ presence [16,17]. It is likely that socioeconomic factors linked to the locality affect the abundance and diversity of breeding containers. However, few studies have been conducted in the same area on socioeconomic and demographic aspects, as well as on knowledge, attitudes, and practices regarding dengue and its vector.

While vector control is jeopardized by widespread resistance to insecticides in *Ae. aegypti* populations throughout Africa, community-based larval source management is emerging as a sustainable control strategy [18]; however, this remains dependent also on the population’s knowledge of dengue and its vector.

Several KAP (Knowledge, Attitudes and Practices) studies on dengue and its vector have been conducted in South America and Asia, but remain a critical gap in research within Africa. These data are crucial for informing the development of effective dengue communication and control strategies. Research in India and Sri Lanka has revealed low levels of knowledge and awareness regarding dengue, its transmission, and preventive measures [19,20]. A multi-country investigation in urban and peri-urban areas of Asia, as well as a study in Bangladesh, found that participants exhibited inadequate knowledge of dengue vector breeding sites and mosquito immature stages [16,17].

One of the most relevant investigations on the population vulnerability to dengue in Ouagadougou by Tougma [21] showed that only 65% of the respondents had heard of dengue and 62% were aware that the disease is transmitted by a mosquito vector. In East Africa, specifically in Ethiopia, a KAP study on dengue showed that a significant proportion (95.1%) of the population had heard of dengue [22]. However, people’s understanding of dengue transmission, its vector, and prevention methods remain inadequate. Additionally, a KAP study conducted in the same country revealed that even among healthcare workers, the level of knowledge about dengue transmission mode and vectors remains limited [23].

Our study aimed to address this knowledge gap for Burkina Faso and was conducted in three Health Districts (HDs) of Ouagadougou to provide an overview of the socio-economic, demographic, and entomological factors affecting dengue epidemiology, its vector and its transmission in the city. More specifically, it maps out (i) the population’s knowledge of dengue and the dengue vector; (ii) the population’s reported practices and attitudes towards dengue, its transmission, and its vector; (iii) the associated factors involved in the knowledge, practices, and proliferation of *Ae. aegypti* and transmission of dengue in the study area; (iv) the types of potential larval containers and their prevalence in the HDs; and, finally (v), the Stegomyia indices (Stegomyia refers to a subgenus of Aedes that includes *Ae aegypti* and *Ae albopictus*) in the study area.

## Method

### Study area and period

A survey was conducted in August 2021, coinciding with the period of high mosquito density, in three out of five HDs in Ouagadougou (N: 12° 22’ 07.64”, W: 001° 31’ 35.63”, Figure 1). The three HDs, Baskuy, Bogodogo, and Nongremassom, are situated in the central, northern, and south-eastern parts of the city, and encompass urban and peri-urban areas.

### Study design and field study

A house-to-house cross-sectional socio-entomological survey was conducted to evaluate the population’s knowledge of dengue and dengue vectors, their perceptions, and attitudes towards the risk of dengue. Data collection took place in both urban and peri-urban areas of the HDs of Bogodogo and Nongremassom, while the HD of Baskuy is entirely urban. We selected 10, 13, and 11 neighbourhoods in the HDs of Baskuy, Bogodogo, and Nongremassom, respectively, based on the geographical extent of the HDs. In each neighbourhood, the characteristics of the visited houses were also documented. Houses classified as modern type have constructions entirely made of cement or concrete, also referred to as “solid construction” with metallic roof. The traditional type consists of constructions made entirely of mud and mud bricks with mud or straw roof. Houses constructed with both types (cement and mud) and metallic roof are categorized as semi-modern or mixed construction.

In each compound, after a detailed study introduction, a questionnaire was provided to a male or female resident who was at least 11 years old, following the acquisition of informed consent from adult respondents and assent from parents or guardians for minors (ages 11 to 20). Residents under 20 years of age (the required minimum age for consent) were also included in the study to explore the variation in knowledge about dengue and its vectors among different age groups.

The study questionnaire was designed to collect socio-demographic, economic, and entomological information of the household. Knowledge, attitudes, and practices related to dengue and its vector were also addressed by the questionnaire. The knowledge on dengue vectors included a practical exercise with Petri dishes containing adults of *Ae. aegypti*, *Anopheles gambiae*, and *Culex quinquefasciatus*; Falcon tubes (50 mL) containing living larvae and pupae of mosquitoes were also prepared. The exercise involved the morphological identification of adult *Ae. aegypti* among Culex and Anopheles, as well as the recognition of larvae and pupae as mosquito’s immature stages.

Questionnaires about dengue and its vector were organized to allow the questionnaire to progress to the next question or stop, depending on the answer. For example, respondents who had never heard of dengue fever were not asked questions about disease’s symptoms or its vector. Similarly, only respondents aware of dengue as a mosquito-borne disease were questioned about mosquito breeding sites, biting periods, and so on. The responses were converted into quantitative numerical data by scoring one point for a correct answer and zero for an incorrect one [24]. Then, we proceeded to sum up the total score for each individual as the level of knowledge about dengue and its vector.

After administering the questionnaire, we conducted a thorough inspection of the entire compound to identify potential *Ae. aegypti* larval sites. These containers were categorized as handwashing stations (HWS) containers, tires, Barrel/Drums (BD), Buckets/cans/pots (BCP), Water feeder (WF), Small containers (SC) (less than 5 litres), and others. We also documented the presence of *Ae. Aegypti* immatures (larvae and/or pupae) in the positive water-holding containers.

The questionnaire and breeding site record forms were implemented on tablets using the KoBoCollect 2021 software[25].

### Statistical analysis

All data analysis were done in R 3.2.1. Comparison among proportions were conducted using chi-square tests with significance level set at p < 0.05. The results of responses to the questionnaire were expressed as proportions, Odd Ratios (OR) and 95% confidence intervals (CI). Three generalized linear mixed models (glmm) with a negative binomial link function utilising the glmmTMB package [26] were fitted to the data with total number of water containers, number of positive containers, and number of productive containers used as response variables. The predictors included HDs, month of collection, type of house, the level of knowledge of the interviewee, the socio-economic and demographic data (number of residents, the type of construction, annual income, water supply), the HDs, and water container type with day of inspection as random effect.

Simple linear models were fitted to knowledge data with dengue and dengue vector knowledge scores as response variables and HDs, month of collection, type of house, socio-economic and demographic data (number of residents, the type of construction, annual income, water supply), sex and age of respondent, the instruction level and the occupation, the annual income, the house type and their interactions (age*sex, age*instruction level*sex, occupation*instruction level).

Minimal models were selected using a stepwise backward procedure based on the lowest AIC values by removing the factors with the highest p-value in the model. If the removal of a variable resulted in a change in the AIC value of more than 2 and the resulting model remained parsimonious, e.g. after the diagnosis of residuals and overdispersion in the R package DHARMa.

The traditional Stegomyia indices[27,28] were assessed: (i) House Index (HI), calculated as the number of houses (or compounds) with at least one *Ae. aegypti* immature positive container × 100/ number of houses visited; (ii) Container Index (CI), calculated as the number of *Ae. aegypti* positive containers × 100/ number of water-holding containers inspected; (iii) Breteau Index (BI), calculated as the number of *Ae. aegypti* positive containers × 100/ number of houses visited.

### Ethical considerations

This study was approved by the Ethical Research Committee of the Ministry of Health (Deliberation n° 2021-08-206 du 04/08/2021). Signed informed consent or assent (for minors) were obtained from all householders included in the study before introducing the questionnaire and commencing the household entomological inspection.

## Results

### Characteristics of populations, compounds, and study areas

A total of 514 individuals from various compounds were interviewed (S1Table). The age range of the respondents was from 11 to 86 years, with an average of 35.49 years and a median of 31 years across the study area. Among the participants, 226 (43.97%) were male and 288 (56.03%) were female. The average number of inhabitants per household was 9.36 for the entire study area, with a significantly higher density in Baskuy (11.60) compared to the other two HDs (p < 0.05), and a statistically lower density in the peri-urban neighbourhoods (6.28) compared to the urban ones (10.46) (p < 0.05).

In the entire study area, 47.5% (244/514) of respondents had not received education beyond the primary school level, with proportions of 42.5% in urban areas and 63.0% in peri-urban areas. Conversely, the percentage of individuals who had attended secondary school was higher in urban areas (57.5%) compared to peri-urban areas (36.1%) (S1Table).

The respondents’ occupations included housewives, government employees, traders, students, artisans, and others. The proportions varied among HD, but in the entire study area students (25.47%, 131/514), traders (21.60%, 111/514), craftsmen (18.68%, 96/514) and housewives/unemployed (15.56%, 80/514) were the most abundant (S1Table).

The surveyed compounds were classified into three types based on building materials. In the HDs of Baskuy, Bogodogo, and Nongremassom, 9.27%, 17.95%, and 23.81% of the compounds are of the traditional type, entirely made of mud bricks. The majority of compounds in the study area are entirely made of cement (modern) (ranging from 41.07% to 65.56%), followed by those of mixed materials (i.e., mud and cement) (ranging from 24.1% to 35.12%) and traditional ones. In the urban area, a similar trend was observed, with cement-based compounds being the most common, followed by those constructed with mixed materials. In contrast, a reverse trend is observed in the peri-urban zone, where compounds made of mud bricks, mixed, and cement were 57.14%, 31.93%, and 10.92%, respectively (S1Table).

The classification of compounds according to income revealed that more than half of the surveyed compounds in all HDs have an annual income of at least 2,400 USD (approximately 1,200,000 FCFA), except for the Nongremassom HD, where only 39.29% of the compounds fall into this category. In the urban area, 54.68% of the compounds have an annual income of at least 2,400 USD (1,200,000 FCFA), while in the peri-urban area, only 30.25% are in this category (S1Table).

In Baskuy, Bogodogo, and Nongremassom, 81.46% (123/151), 67.18% (131/195), and 60.71% (102/168) of the compounds had access to piped water, respectively, compared to 84.05% (332/395) and 20.17% (24/119) in urban and peri-urban areas (S1Table).

### Knowledge of dengue

The respondents’ knowledge level regarding dengue, including its symptoms and transmission mode, indicated that 94.36% (485/514) were familiar with dengue fever (Table 1). Specifically, 97.35% (147/151), 90.26% (176/195), and 96.43% (162/168) of respondents in the HDs of Baskuy, Bogodogo, and Nongremassom fell into this category. These proportions were found to be statistically different (p = 0.007) (Table 1). When comparing types of settlement, 95.19% (376/395) of urban respondents and 91.60% (109/119) of peri-urban respondents had heard of dengue fever, with no significant difference between these proportions (p = 0.207).

**Table 1.**
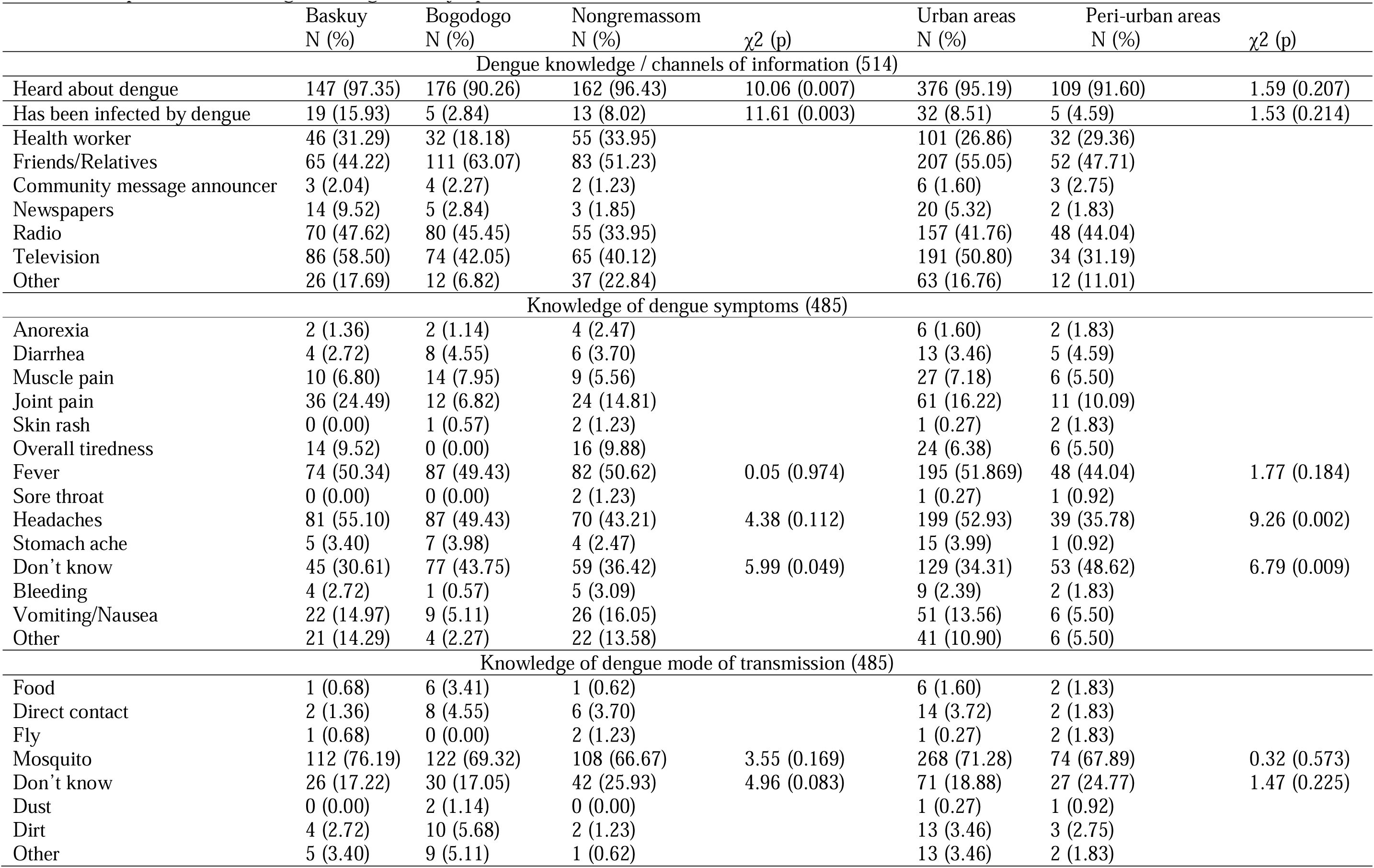
Population knowledge of dengue, its symptoms and its transmission mode.

| | Baskuy<br>N (%) | Bogodogo<br>N (%) | Nongremassom<br>N (%) | $\chi^2$ (p) | Urban areas<br>N (%) | Peri-urban areas<br>N (%) | $\chi^2$ (p) |
| --- | --- | --- | --- | --- | --- | --- | --- |
| Dengue knowledge / channels of information (514) |  |  |  |  |  |  |  |
| Heard about dengue | 147 (97.35) | 176 (90.26) | 162 (96.43) | 10.06 (0.007) | 376 (95.19) | 109 (91.60) | 1.59 (0.207) |
| Has been infected by dengue | 19 (15.93) | 5 (2.84) | 13 (8.02) | 11.61 (0.003) | 32 (8.51) | 5 (4.59) | 1.53 (0.214) |
| Health worker | 46 (31.29) | 32 (18.18) | 55 (33.95) |  | 101 (26.86) | 32 (29.36) |  |
| Friends/Relatives | 65 (44.22) | 111 (63.07) | 83 (51.23) |  | 207 (55.05) | 52 (47.71) |  |
| Community message announcer | 3 (2.04) | 4 (2.27) | 2 (1.23) |  | 6 (1.60) | 3 (2.75) |  |
| Newspapers | 14 (9.52) | 5 (2.84) | 3 (1.85) |  | 20 (5.32) | 2 (1.83) |  |
| Radio | 70 (47.62) | 80 (45.45) | 55 (33.95) |  | 157 (41.76) | 48 (44.04) |  |
| Television | 86 (58.50) | 74 (42.05) | 65 (40.12) |  | 191 (50.80) | 34 (31.19) |  |
| Other | 26 (17.69) | 12 (6.82) | 37 (22.84) |  | 63 (16.76) | 12 (11.01) |  |
| Knowledge of dengue symptoms (485) |  |  |  |  |  |  |  |
| Anorexia | 2 (1.36) | 2 (1.14) | 4 (2.47) |  | 6 (1.60) | 2 (1.83) |  |
| Diarrhea | 4 (2.72) | 8 (4.55) | 6 (3.70) |  | 13 (3.46) | 5 (4.59) |  |
| Muscle pain | 10 (6.80) | 14 (7.95) | 9 (5.56) |  | 27 (7.18) | 6 (5.50) |  |
| Joint pain | 36 (24.49) | 12 (6.82) | 24 (14.81) |  | 61 (16.22) | 11 (10.09) |  |
| Skin rash | 0 (0.00) | 1 (0.57) | 2 (1.23) |  | 1 (0.27) | 2 (1.83) |  |
| Overall tiredness | 14 (9.52) | 0 (0.00) | 16 (9.88) |  | 24 (6.38) | 6 (5.50) |  |
| Fever | 74 (50.34) | 87 (49.43) | 82 (50.62) | 0.05 (0.974) | 195 (51.869) | 48 (44.04) | 1.77 (0.184) |
| Sore throat | 0 (0.00) | 0 (0.00) | 2 (1.23) |  | 1 (0.27) | 1 (0.92) |  |
| Headaches | 81 (55.10) | 87 (49.43) | 70 (43.21) | 4.38 (0.112) | 199 (52.93) | 39 (35.78) | 9.26 (0.002) |
| Stomach ache | 5 (3.40) | 7 (3.98) | 4 (2.47) |  | 15 (3.99) | 1 (0.92) |  |
| Don't know | 45 (30.61) | 77 (43.75) | 59 (36.42) | 5.99 (0.049) | 129 (34.31) | 53 (48.62) | 6.79 (0.009) |
| Bleeding | 4 (2.72) | 1 (0.57) | 5 (3.09) |  | 9 (2.39) | 2 (1.83) |  |
| Vomiting/Nausea | 22 (14.97) | 9 (5.11) | 26 (16.05) |  | 51 (13.56) | 6 (5.50) |  |
| Other | 21 (14.29) | 4 (2.27) | 22 (13.58) |  | 41 (10.90) | 6 (5.50) |  |
| Knowledge of dengue mode of transmission (485) |  |  |  |  |  |  |  |
| Food | 1 (0.68) | 6 (3.41) | 1 (0.62) |  | 6 (1.60) | 2 (1.83) |  |
| Direct contact | 2 (1.36) | 8 (4.55) | 6 (3.70) |  | 14 (3.72) | 2 (1.83) |  |
| Fly | 1 (0.68) | 0 (0.00) | 2 (1.23) |  | 1 (0.27) | 2 (1.83) |  |
| Mosquito | 112 (76.19) | 122 (69.32) | 108 (66.67) | 3.55 (0.169) | 268 (71.28) | 74 (67.89) | 0.32 (0.573) |
| Don't know | 26 (17.22) | 30 (17.05) | 42 (25.93) | 4.96 (0.083) | 71 (18.88) | 27 (24.77) | 1.47 (0.225) |
| Dust | 0 (0.00) | 2 (1.14) | 0 (0.00) |  | 1 (0.27) | 1 (0.92) |  |
| Dirt | 4 (2.72) | 10 (5.68) | 2 (1.23) |  | 13 (3.46) | 3 (2.75) |  |
| Other | 5 (3.40) | 9 (5.11) | 1 (0.62) |  | 13 (3.46) | 2 (1.83) |  |

Most respondents heard about dengue fever through radio (42.27%), television (46.39%), and friends or relatives (53.40%). Among them, 12.93% (19/147), 2.84% (5/176), and 8.02% (13/162) of the respondents in the HDs of Baskuy, Bogodogo, and Nongremassom reported having contracted dengue, showing a significant difference in proportions (p = 0.003) (Table 1). In the urban and peri-urban categories, 8.51% (32/376) and 4.59% (5/109) of the respondents declared having contracted dengue, with no significant difference between the two areas (p = 0.214) (Table 1). Regarding dengue symptoms, roughly half of those aware of dengue (50.34% in Baskuy, 49.64% in Bogodogo, and 50.62% in Nongremassom) linked it to fever. Additionally, 55.10%, 49.43%, and 43.21% of the respondents in Baskuy, Bogodogo, and Nongremassom, respectively mentioned headaches as a symptom. Across the three HDs, 30.61% (45/147), 43.75% (77/176), and 36.42% (59/162) of individuals from Baskuy, Bogodogo, and Nongremassom, respectively, did not know any dengue symptoms, with a significant difference among HDs (p < 0.05) (Table 1). These proportions were 34.31% (129/376) and 48.62% (53/109) in the urban and peri-urban areas, respectively, with a significant difference between the two areas (p = 0.009) (Table 1). In the overall study area, 37.53% (182/485) of respondents aware of dengue were unfamiliar with any symptoms of the disease.

Knowledge of the dengue vectors varied among respondents in different health districts. In Baskuy, 76.19% (112/147) knew that dengue transmission occurred through a mosquito bite, while in Bogodogo and Nongremassom, the percentages were 69.32% (122/176) and 66.67% (108/162) respectively. Comparatively, 71.28% (268/376) of urban respondents and 67.89% (74/109) of peri-urban respondents were aware of the mosquito vector. Overall, 70.52% (342/485) of all respondents in the study area understood that dengue is transmitted by mosquitoes (Table 1).

### Knowledge of the dengue vectors

The questions on the knowledge of the vector were selectively submitted to those who knew that dengue is transmitted by a mosquito and, of them, 80.12% (274/342) knew that the dengue vector needs water to breed (Table 2). This proportion varied between HDs, with 91.80% (112/122) in Bogodogo 73.21% (82/112) to 74.07% (80/108) in Baskuy and Nongremassom respectively with a significant difference between the HDs (P < 0.001). The difference between proportions from the urban and the peri-urban areas respectively of 79.10% (212/268) and 83.78% (62/74) was not statistically significant (p = 0.466) (Table 2).

**Table 2.** Population knowledge and pratices about the dengue vector, its breeding places, its biting activity periods, its morphological aspects (adult) and ability to recognize mosquito larvae (of mosquitoes in general)

| | Baskuy<br>N (%) | Bogodogo<br>N (%) | Nongremassom<br>N (%) | $\chi^2$ (p) | Urban areas<br>N (%) | Peri-urban areas<br>N (%) | $\chi^2$ (p) |
| --- | --- | --- | --- | --- | --- | --- | --- |
| Knowledge of the breeding places of dengue mosquitoes (342) |  |  |  |  |  |  |  |
| Does mosquito need water to breed? | 82 (73.21) | 112 (91.80) | 80 (74.07) | 16.26 (<0.001) | 212 (79.10) | 62 (83.78) | 0.53 (0.466) |
| Knowledge on dengue mosquitoes breeding site types (274) |  |  |  |  |  |  |  |
| Animal water feeders | 5 (6.10) | 5 (4.46) | 5 (6.25) |  | 9 (4.25) | 6 (9.68) |  |
| Drainage channels | 14 (17.07) | 20 (17.86) | 14 (17.25) |  | 41 (19.34) | 7 (11.29) |  |
| Waste | 24 (29.27) | 55 (49.11) | 40 (50.) |  | 89 (41.98) | 30 (48.39) |  |
| Stagnant clean water | 38 (46.34) | 61 (54.46) | 31 (38.75) |  | 102 (48.11) | 28 (45.16) |  |
| Dirty water | 65 (79.27) | 107 (95.54) | 57 (71.25) |  | 173 (81.60) | 56 (90.32) |  |
| Don't know | 1 (1.22) | 0 (0) | 1 (1.25) |  | 2 (0.94) | 0 (0.0) |  |
| Tires | 3 (3.66) | 9 (8.06) | 8 (10) |  | 16 (67.55) | 4 (6.45) |  |
| Flower pot | 6 (7.32) | 4 (3.57) | 9 (11.25) |  | 15 (7.08) | 4 (6.45) |  |
| Water storage containers | 18 (21.95) | 57 (50.89) | 22 (27.50) |  | 79 (37.26) | 18 (29.03) |  |
| Other | 17 (20.73) | 4 (3.57) | 20 (25.00) |  | 37 (17.45) | 4 (6.45) |  |
| Knowledge of Dengue Mosquito Biting Periods (342) |  |  |  |  |  |  |  |
| Sunrise | 9 (8.04) | 10 (8.20) | 13 (12.04) | 1.34 (0.512) | 25 (9.33) | 7 (9.46) | 0.00 (0.999) |
| Sunset | 21 (18.75) | 16 (13.11) | 18 (16.67) | 1.41 (0.493) | 43 (16.04) | 12 (16.22) | 0.00 (0.999) |
| Daytime/Sunrise/Sunset | 83 (74.11) | 104 (85.25) | 72 (66.67) | 11.00 (0.004) | 202 (75.37) | 57 (77.03) | 0.02 (0.888) |
| Nighttime | 30 (26.79) | 66 (54.10) | 37 (34.26) |  | 92 (34.33) | 41 (55.41) |  |
| Daytime | 69 (61.61) | 93 (76.23) | 61 (56.48) |  | 171 (63.81) | 52 (70.27) |  |
| Don't know | 20 (17.86) | 10 (8.20) | 18 (16.67) |  | 41 (15.30) | 7 (9.46) |  |
| Knowledge of the characteristics of the dengue vector mosquito (342) |  |  |  |  |  |  |  |
| Black | 10 (8.93) | 38 (31.15) | 8 (7.41) |  | 35 (13.06) | 21 (28.38) |  |
| Brown | 0 (0.00) | 0 (0.00) | 1 (0.93) |  | 1 (0.37) | 0 (0.00) |  |
| Black with white spots | 45 (40.18) | 29 (23.77) | 35 (32.41) | 7.26 (0.027) | 95 (35.45) | 15 (20.27) | 5.45 (0.020) |
| Don't know | 56 (50.00) | 55 (45.08) | 64 (59.26) |  | 137 (51.12) | 38 (51.35) |  |
| Recognizes dengue mosquito vector | 45 (40.18) | 29 (23.77) | 35 (32.41) | 7.26 (0.027) | 95 (35.45) | 15 (20.27) | 5.45 (0.020) |
| Recognizes mosquito larvae (in general) | 18 (16.07) | 13 (10.66) | 18 (16.67) | 2.10 (0.350) | 42 (15.67) | 7 (9.46) | 1.35 (0.245) |

Regarding dengue vector breeding places, the respondents cited clean stagnant water containers (47.45%, 130/274), water storage containers (35.40%, 97/274), tires (7.30%, 20/274), flower pots (6.93%, 19/274), and animal water feeders (5.47%, 15/274). Many respondents (85.58%, 229/274) across the HDs also mentioned dirty water (Table 2).

With regards to the dengue vector biting period, 74.11% (83/112), 85.25% (104/122), and 66.67% (72/108) in the HDs of Baskuy, Bogodogo, and Nongremassom, respectively, associated the dengue vector with daytime biting activity (p = 0.004). In comparison, these proportions in the urban and peri-urban areas were 75.37% (202/268) and 77.03% (57/74) respectively, with no significant differences (p = 0.888) (Table 2).

Regarding morphological recognition of the adult mosquito of *Ae. aegypti*, 40.17% (45/112), 23.77% (29/122) and 32.41% (35/108) of the respondents in the HDs of Baskuy, Bogodogo and Nongremassom, recognised the adult stage of *Ae. aegypti*, among *An. gambiae* and *Cx. quinquefasciatus* adult mosquitoes, with significant differences (p = 0.027) between HDs (Table 2). In the urban and peri-urban areas, the proportion of those capable of identifying *Ae. aegypti* from the two other species was 35.07% (94/268) and 21.62% (16/74) respectively, with a statistically significant difference between urban and peri-urban areas (p = 0.024). Across the study area, 32.16% (110/342) of respondents were able to morphologically identify the dengue vector among other non-Aedes mosquito species (Table 2).

In terms of recognition of mosquito larvae in general, fewer than 15% of respondents in the study area could make a link between larvae and adult mosquito in general (Table 2). Only 16.07% (18/112), 10.66% (13/122), and 16.67% (18/108) of respondents in the HDs of Baskuy, Bogodogo, and Nongremassom, respectively, could recognize a mosquito larva, compared to 15.42% (42/268) and 9.46% (7/74) in the urban and peri-urban areas, respectively, with no significant statistical difference either among HDs (p = 0.350) or among urban and peri-urban areas (p = 0.245) (Table 2).

### People’s practices and attitudes regarding dengue transmission and prevention

#### Practices that may increase mosquito breeding or biting

Practices that may create favourable conditions for dengue vector mosquito breeding and promote human-vector contact are outlined in Table 3. At health districts (HDs) level as well as in urban *versus* peri-urban areas, rainwater harvesting containers were present in over 90% of households (Table 3A). Some of these containers already contained water and were potential larval sites for Aedes mosquitoes. In the three HDs, 17.22% to 38.10% of the compounds received water from public fountains, with a statistically significant difference between these proportions in the HDs (p < 0.001). The difference was particularly marked between compounds in the urban and peri-urban areas that relied on water from public fountains, 14.18% (56/395) and 79.83% (95/119), respectively, also, showing a significant difference between these proportions (p < 0.001).

**Table 3:** Population practices and attitudes that may have an effect on dengue transmission in the study area.

| | Baskuy<br>N (%) | Bogodogo<br>N (%) | Nongremassom<br>N (%) | $\chi^2$ (p) | Urban areas<br>N (%) | Peri-urban<br>areas N (%) | $\chi^2$ (p) |
| --- | --- | --- | --- | --- | --- | --- | --- |
| A- Presence of potential *Aedes aegypti* breeding sites (514) |  |  |  |  |  |  |  |
| Presence of containers | 138 (91.39) | 181 (92.82) | 152 (90.48) | 0.66 (0.718) | 359 (90.87) | 112 (94.12) | 0.86 (0.354) |
| Presence of containers with water | 129 (85.43) | 174 (89.23) | 145 (86.31) | 0.13 (0.533) | 341 (86.33) | 107 (89.92) | 0.76 (0.384) |
| Presence of containers / *Aedes aegypti* pupae | 74 (49.01) | 59 (30.26) | 109 (64.88) | 43.74 (<0.001) | 196 (49.62) | 46 (38.66) | 3.98 (0.045) |
| B- Practices that favour mosquito breeding or biting (514) |  |  |  |  |  |  |  |
| Water supply at public fountain | 26 (17.22) | 61 (31.28) | 64 (38.10) | 17.26 (<0.001) | 56 (14.18) | 95 (79.83) | 186.85 (0.001) |
| Presence of open storage container | 80 (52.98) | 160 (82.05) | 122 (72.62) | 35.11 (<0.001) | 262 (66.33) | 100 (84.03) | 12.93 (<0.001) |
| Water storage practice | 61 (40.40) | 142 (72.82) | 82 (48.81) | 40.67 (<0.001) | 189 (47.85) | 96 (80.67) | 38.57 (<0.001) |
| Presence of waste that can hold water | 89 (58.94) | 164 (84.10) | 92 (54.76) | 41.69 (0.001) | 251 (63.54) | 94 (78.99) | 9.20 (0.002) |
| Do not overturn water from unused containers | 85 (56.29) | 170 (87.18) | 122 (72.62) | 41.60 (0.001) | 297 (75.19) | 80 (67.23) | 2.57 (0.109) |
| Absence of curtains/doors & windows | 5 (3.31) | 4 (2.05) | 11 (6.55) | - | 13 (3.29) | 7 (5.88) | - |
| Absence of mosquito screens | 78 (51.66) | 154 (78.97) | 108 (64.29) | 28.75 (0.001) | 234 (59.24) | 106 (89.08) | 35.03 (<0.001) |
| C- Practices to protect against mosquito bite (514) |  |  |  |  |  |  |  |
| Bed net use/ night-time | 110 (72.85) | 178(91.28) | 136 (80.95) | 20.43 (0.001) | 321 (81.27) | 103 (86.55) | 1.42 (0.233) |
| Bed net use/ daytime | 27 (17.88) | 28 (14.36) | 32 (19.05) | 1.55 (0.461) | 66 (16.71) | 21 (17.65) | 0.01 (0.921) |
| Use of anti-mosquito lotion | 38 (25.16) | 15 (7.69) | 54 (32.14) | 35.18 (<0.001) | 99 (25.06) | 8 (6.72) | 17.53 (<0.001) |
| Use of anti-mosquito fumigant | 116 (76.82) | 178 (91.28) | 138 (82.14) | 13.95 (0.001) | 322 (82.52) | 110 (92.43) | 7.34 (0.007) |
| Use of anti-mosquito spray | 73 (48.34) | 103 (52.88) | 64 (38.10) | 8.09 (0.022) | 187 (47.34) | 53 (43.54) | 0.19 (0.663) |
| Use of fan | 3 (1.99) | 1 (0.51) | 9 (5.36) | - | 012 (3.04) | 1 (0.84) | - |
| Others anti-mosquito means | 5 (3.31) | 2(1.03) | 20 (11.90) | - | 22 (5.57) | 5 (4.20) | - |
| D- Respondents' attitudes toward mosquito bite and dengue prevention methods (485) |  |  |  |  |  |  |  |
| Destroy larvae breeding sites | 27 (18.37) | 22 (12.50) | 45 (27.78) | 12.74 (0.002) | 83 (22.07) | 11 (10.09) | 7.71 (0.006) |
| Empty containers with water | 63 (42.86) | 75 (42.61) | 81 (50.00) | 3.58 (0.167) | 187 (49.73) | 32 (29.36) | 14.82 (<0.001) |
| Cover water storage containers | 21 (14.29) | 34 (19.32) | 17 (10.49) | 4.01 (0.135) | 62 (16.49) | 10 (9.17) | 3.02 (0.082) |
| Sleep under a mosquito bed net | 85 (57.82) | 163 (92.61) | 118 (72.84) | 53.27 (<0.001) | 278 (73.94) | 88 (80.73) | 1.76 (0.185) |
| Use insecticide/repellent | 59 (40.13) | 132 (75.00) | 64 (39.51) | 55.72 (<0.001) | 196 (49.62) | 59 (45.38) | 0.07 (0.795) |
| Don't know | 20 (13.61) | 5 (2.84) | 13 (8.02) | - | 30 (7.98) | 8 (7.34) | - |
| Cleanliness | 25 (17.01) | 7 (3.98) | 13 (8.02) | - | 36 (9.57) | 9 (8.26) | - |
| Long-sleeved clothes | 13 (8.84) | 2 (1.14) | 10 (6.17) | - | 24 (6.38) | 1 (0.92) | - |
| Other | 6 (4.08) | 2 (1.14) | 23 (14.20) | - | 26 (6.91) | 5 (4.59) | - |

Water storage for later use was relatively common, as noted among 40.40% to 72.82% of the compounds in the HDs (Table 3B), with statistically significant variation (p < 0.001); this proportion increased to 47.85% (189/395) and 80.67% (96/119) in the urban and peri-urban areas, with statistical significance (p < 0.001), making a total of 55.45% in the entire study area. The presence of uncovered water storage containers was reported in 70.43% (362/514) of the compounds.

It was observed that in more than half of the compounds in all HDs (ranging from 54.76% to 84.10%), waste materials were present, which could accumulate rainwater and potentially serve as larval sites for mosquitoes. Furthermore, the lack of a habit among the population to empty water from unused containers (during the rainy season) that could become larval sites was noted in 73.35% of compounds. Moreover, 3.89% and 66.15% of compounds had no curtains on doors and/or windows, and no mosquito screens on house windows, respectively.

#### Practices to protect against mosquito bites

Protective practices against dengue vector bites are detailed in Table 3C. Specifically, 72.85% (110/151), 91.28% (178/195), and 80.95% (136/168) of respondents in the HDs of Baskuy, Bogodogo, and Nongremassom respectively use bed nets at night. A significant difference was noted between HDs (p < 0.001). However, 81.27% (321/395) and 86.55% (103/119) of respondents in urban and peri-urban areas use bed nets at night, with no statistically significant difference observed (p = 0.232). In contrast, less than 20% of individuals in all HDs, urban, and peri-urban areas declared using bed nets during daytime.

Several other practices are utilized by respondents to protect themselves from mosquito bites, including anti-mosquito fumigants, anti-mosquito bombs, anti-mosquito lotions, fans, and others. Mosquito fumigants and mosquito bombs are the most commonly used, with 84.8% (432/514) and 46.69% (240/514) of respondents employing these practices in the entire study area, respectively.

#### Respondents’ attitudes on dengue and mosquito bites prevention

Attitudes towards protection from dengue vector bites are outlined in Table 3D. In the Health Districts of Baskuy, Bogodogo, and Nongremassom, 57.82% (85/147), 92.61% (163/176), and 72.84% (118/162) of respondents believed that sleeping under a mosquito bed net could prevent mosquito bites and dengue. The difference in these percentages across the HDs is statistically significant (p < 0.001). However, there was no statistically significant distinction between urban and peri-urban areas, where 73.94% (278/376) and 80.73% (88/109) respectively, viewed bed nets as a form of protection against mosquitoes (p = 0.184).

In the entire study area, some respondents reported preventive attitudes against dengue and its vector, including the use of insecticides and repellents (52.58%; 255/485), emptying small water-holding containers (45.15%; 219/485), destroying mosquito breeding sites (19.38%; 94/485), and covering water-holding containers (14.85%; 72/485). Conversely, other respondents believed that maintaining a clean environment (9.28%, 45/485) and wearing long-sleeved clothing (5.15%, 25/485) were also protective measures against dengue and its vector.

### Factors associated with knowledge of dengue, its symptoms and transmission

Levels of knowledge and awareness about dengue, its symptoms, and transmission modes were evaluated among individuals in the study area to explore their association with socio-demographic factors including education, gender, age, respondent status, and residential area (urban or peri-urban) using odds ratio (OR) analyses (Table 4).

**Table 4:** Knowledge of dengue, symptoms, transmission and associated factors.

| Characteristics |  | Don't know about dengue |  |  |  |  |
| --- | --- | --- | --- | --- | --- | --- |
|  |  | Yes | No | OR | 95% CI | χ2 (p) |
| Education level | < Primary | 8 | 95 | 1.56 | 0.67 - 3.64 | 1.04 (0.299) |
|  | Primary and more | 21 | 390 |  |  |  |
| Areas | Peri-urban | 10 | 109 | 1.82 | 0.82 - 4.02 | 1.47 (0.141) |
|  | Urban | 19 | 376 |  |  |  |
| Gender | Female | 22 | 266 | 2.59 | 1.08 - 6.18 | 2.14 (0.032) |
|  | Male | 7 | 219 |  |  |  |
| Age (years) | [11; 30] | 23 | 226 | 4.40 | 1.75 - 11.00 | 3.17 (0.001) |
|  | [31; 88] | 6 | 259 |  |  |  |
| Characteristics |  | Knowledge of dengue major symptoms (Fever and headaches) |  |  |  |  |

| | | Yes | No | OR | 95% CI | $\chi^2$ (p) |
| --- | --- | --- | --- | --- | --- | --- |
| Education level | < Primary | 40 | 55 | 0.43 | 0.26 - 0.68 | 3.67 (< 0.001) |
|  | Primary and more | 246 | 144 |  |  |  |
| Areas | Peri-urban | 56 | 53 | 0.67 | 0.43 - 1.04 | 1.82 (0.068) |
|  | Urban | 230 | 146 |  |  |  |
| Gender | Female | 145 | 121 | 0.66 | 0.45 - 0.96 | 2.19 (0.028) |
|  | Male | 141 | 78 |  |  |  |
| Age (years) | [11; 30] | 132 | 94 | 0.96 | 0.66 - 1.38 | 0.23 (0.814) |
|  | [31; 88] | 154 | 105 |  |  |  |
| Characteristics |  | Don't know any symptoms |  |  |  |  |
| | | Yes | No | OR | 95% CI | $\chi^2$ (p) |
| Education level | < Primary | 54 | 41 | 2.70 | 1.70 - 4.27 | 4.25 (< 0.001) |
|  | Primary and more | 128 | 262 |  |  |  |
| Areas | Peri-urban | 52 | 57 | 1.73 | 1.12 - 2.66 | 2.48 (0.013) |
|  | Urban | 130 | 246 |  |  |  |
| Gender | Female | 114 | 152 | 1.67 | 1.14 - 2.43 | 2.66 (0.008) |
|  | Male | 68 | 151 |  |  |  |
| Age (years) | [11; 30] | 82 | 143 | 0.92 | 0.63 - 1.33 | 0.46 (0.647) |
|  | [31; 88] | 100 | 160 |  |  |  |
| Characteristics |  | Know dengue transmission mode |  |  |  |  |
| | | Yes | No | OR | 95% CI | $\chi^2$ (p) |
| Education level | < Primary | 54 | 41 | 0.47 | 0.29 - 0.75 | 3.22 (0.001) |
|  | Primary and more | 288 | 102 |  |  |  |
| Areas | Peri-urban | 74 | 35 | 0.85 | 0.53 - 1.35 | 0.68 (0.495) |
|  | Urban | 268 | 108 |  |  |  |
| Gender | Female | 175 | 91 | 0.60 | 0.40 - 0.90 | 2.51 (0.012) |
|  | Male | 167 | 52 |  |  |  |
| Age (years) | [11; 30] | 154 | 72 | 0.81 | 0.54 - 1.20 | 1.07 (0.284) |
|  | [31; 88] | 188 | 71 |  |  |  |

Gender and age were key factors associated with different degrees of knowledge of dengue symptoms and transmission. The results indicated that a lack of knowledge about dengue fever, specifically never having heard of it, was associated with the respondent’s gender and age. Female respondents (OR = 2.59; 95% CI = 1.08 - 6.18; p = 0.032) and those younger than 30 years old (OR = 4.40; 95% CI = 1.75 - 11.00; p = 0.001) were more likely to have lower knowledge about dengue fever.

When it comes to understanding the main symptoms of dengue, such as fever and headache, individuals with an education level below primary school (OR = 0.43; 95% CI = 0.26 - 0.68; p < 0.001) and females (OR = 0.66; 95% CI = 0.45 - 0.96; p = 0.028) were found to be less likely to be aware of these symptoms.

Factors such as having an educational level below primary school (OR = 2.70; 95% CI = 1.70 - 4.27; p < 0.001), residing in a peri-urban area (OR = 1.73; 95% CI = 1.12 - 2.66; p = 0.013), and being female (OR = 1.67; 95% CI = 1.14 - 2.43; p = 0.008) were associated with a lack of awareness of all dengue symptoms. Knowledge of dengue transmission modes was linked to the educational level and gender of the respondent. Individuals with less than a primary education (OR = 0.47; 95% CI = 0.29 - 0.75; p = 0.001) and females (OR = 0.60; 95% CI = 0.40 - 0.90; p = 0.012) were less likely to recognize that dengue is a mosquito-borne disease.

According to the glmm (Table 5), general knowledge of dengue fever was positively correlated with respondent age and education level. Populations from Bogodogo have a better knowledge of dengue compared to the reference locality (Baskuy) and Nongremasson. Men appear to have lower knowledge of dengue than women, but all occupational categories show better knowledge of dengue than housewives who also belong to the less educated category. The interaction between sex and instruction level is negatively associated with dengue knowledge for male x no formal schooling. Government employees exhibited better general knowledge of dengue fever compared to other occupations. Knowledge of dengue is correlated to education level with people with university education having better knowledge of dengue. Additionally, the economic status and type of construction of the respondents’ compound were found to be associated with the level of general knowledge about dengue fever.

**Table 5:**
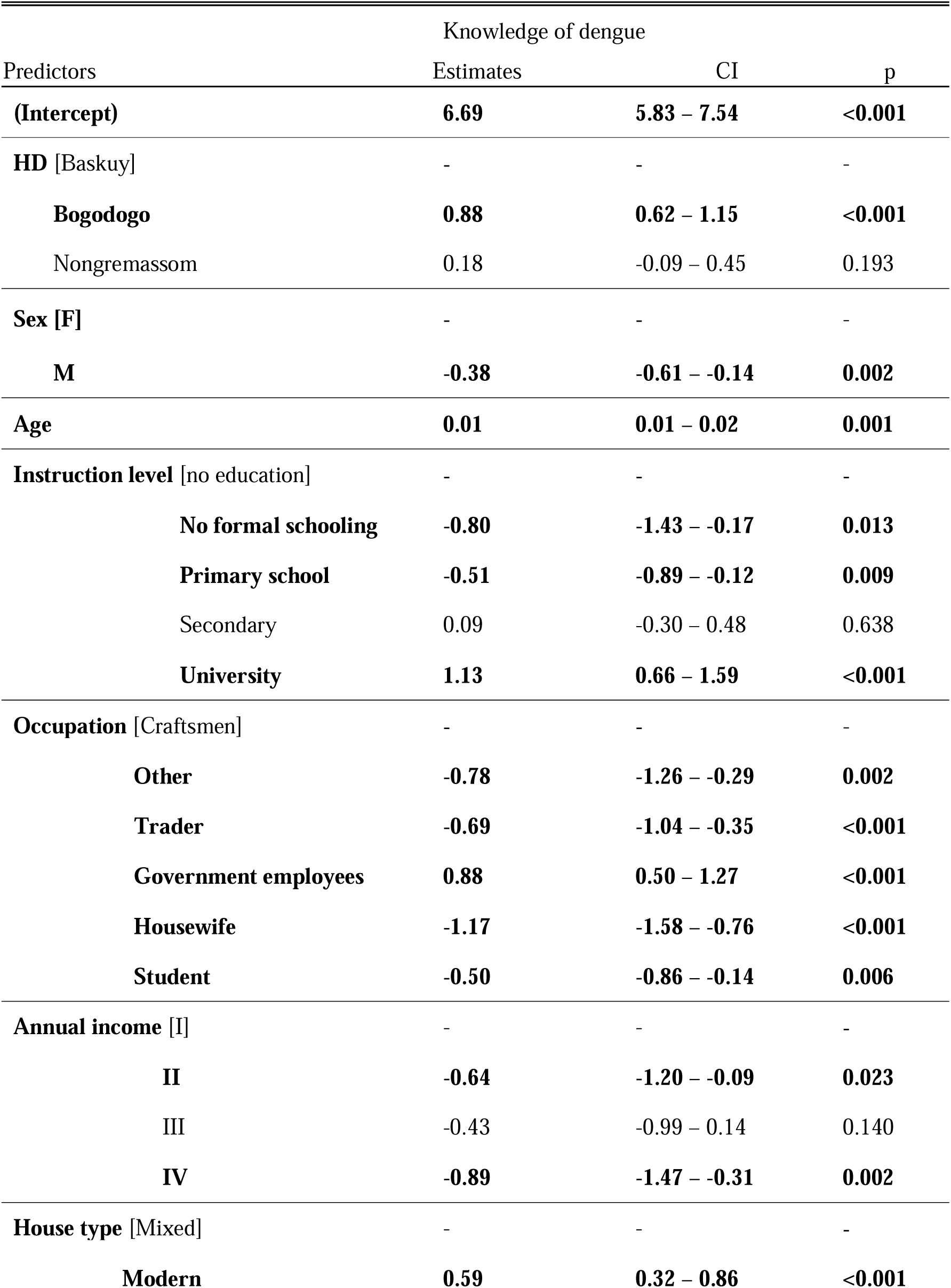
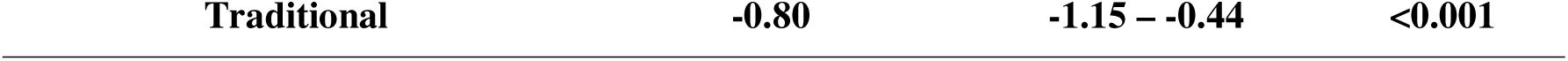
Associations between the dengue knowledge level and different characteristics of the visited compound, respondents’ socioeconomic status and health district following a Generalized Linear Model analysis (glm). Significant predictors are bolded.

### Factors associated with dengue vector knowledge and breeding sites’ presence

Following the OR analysis, Table 6 showed associations between knowledge of the dengue vector (biting periods, mosquito larvae) and socio-demographic characteristics such as the respondent’s age and residential area (urban or peri-urban). Individuals residing in peri-urban areas were more likely to be unaware of the dengue vector biting period (OR = 1.87; 95% CI = 1.09 - 3.20; p = 0.021) compared to those in urban areas. Also, compound which annual income < 1,200,000 FCFA are more likely to be infested with Aedes positive container (OR = 0.53; 95% CI = 0.36 - 0.76; p < 0.001). Respondents aged 11-30 were less likely to be aware of mosquito larvae (OR = 0.39; 95% CI = 0.19 - 0.76; p = 0.006) than older respondents.

**Table 6:**
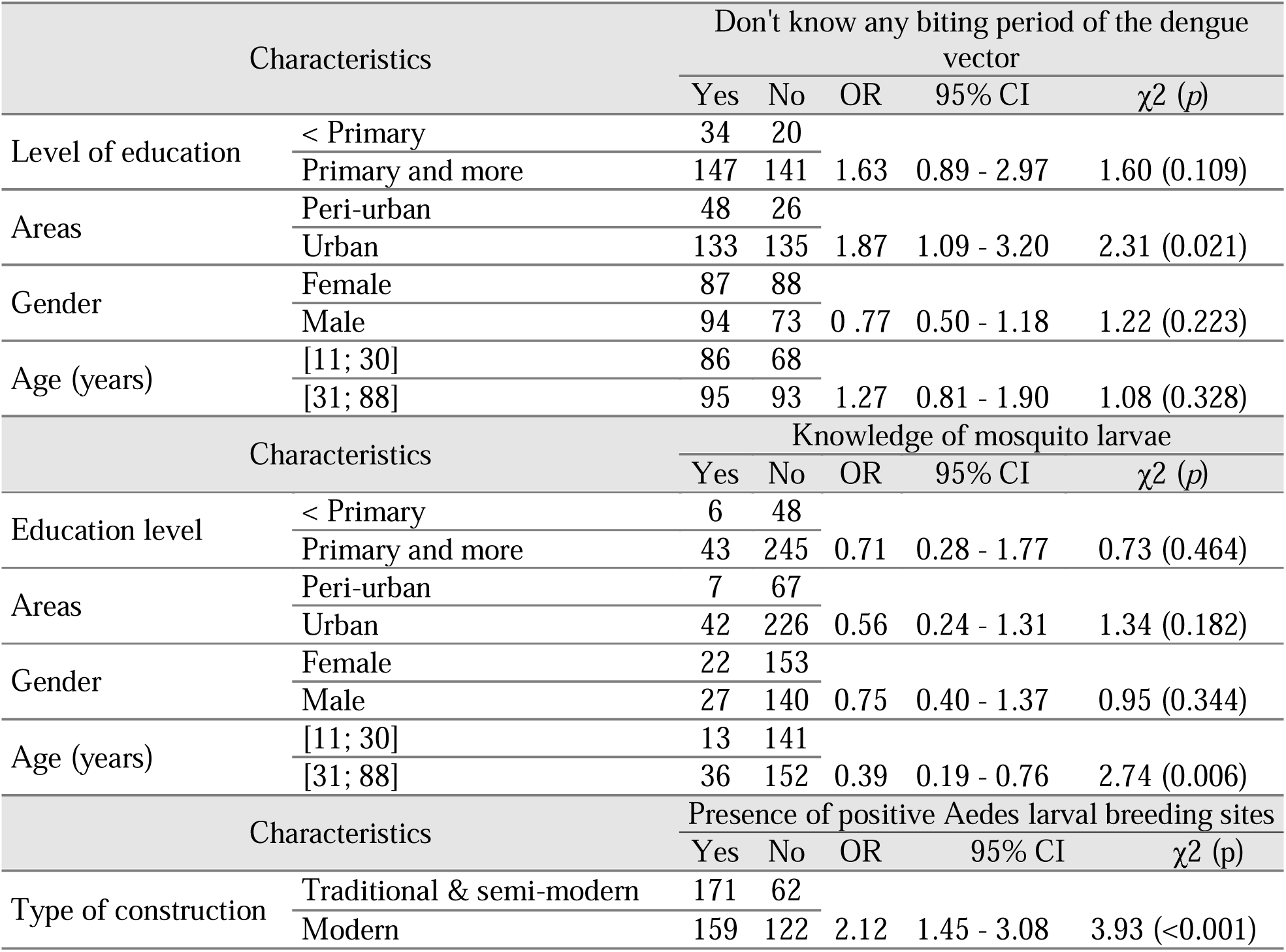
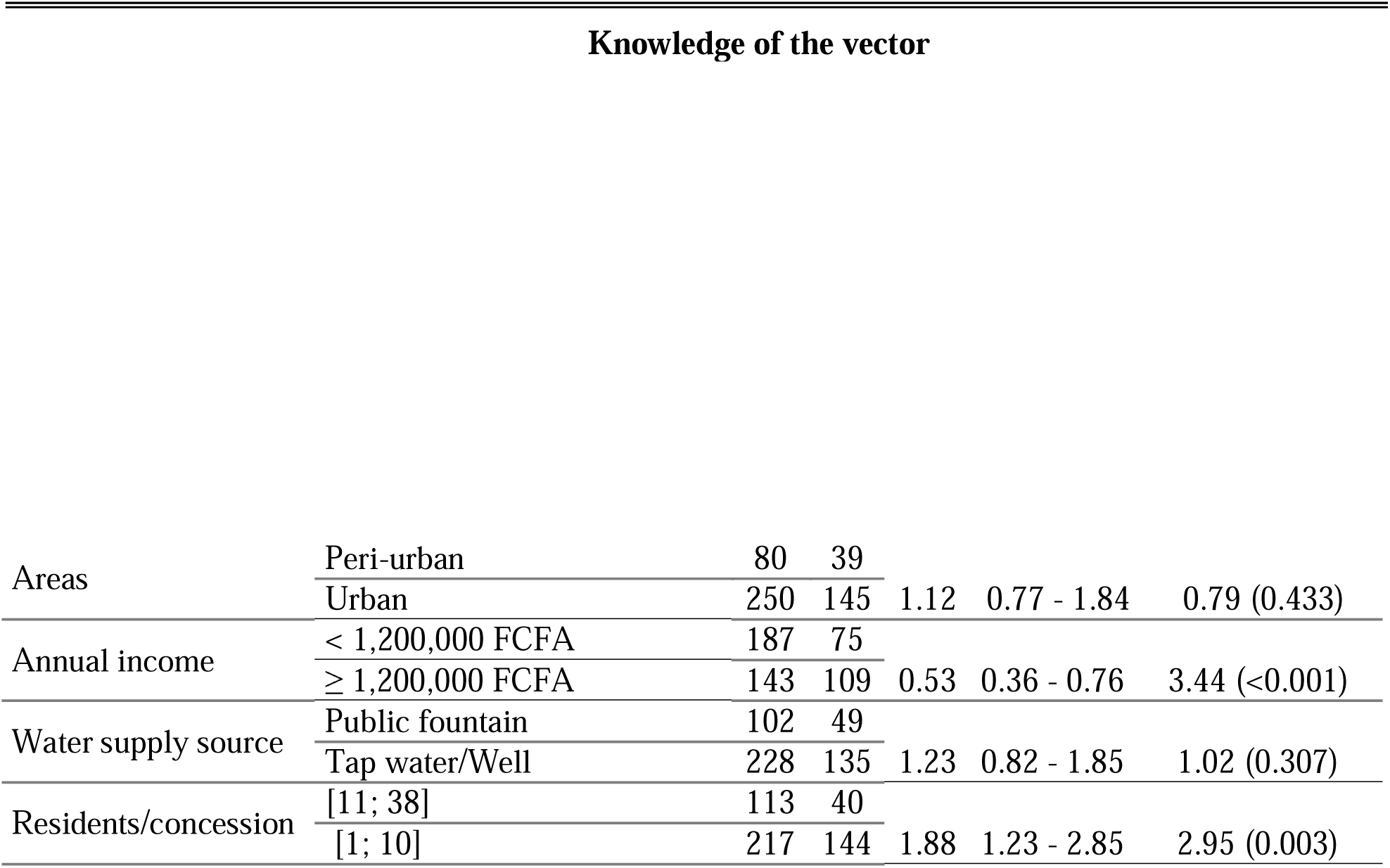
Knowledge of dengue vector and presence of positive Aedes larval sites and their associated factors.

| Characteristics |  | Don't know any biting period of the dengue vector |  |  |  |  |
| --- | --- | --- | --- | --- | --- | --- |
| | | Yes | No | OR | 95% CI | $\chi^2$ (p) |
| Level of education | < Primary | 34 | 20 |  |  |  |
|  | Primary and more | 147 | 141 | 1.63 | 0.89 - 2.97 | 1.60 (0.109) |
| Areas | Peri-urban | 48 | 26 |  |  |  |
|  | Urban | 133 | 135 | 1.87 | 1.09 - 3.20 | 2.31 (0.021) |
| Gender | Female | 87 | 88 |  |  |  |
|  | Male | 94 | 73 | 0.77 | 0.50 - 1.18 | 1.22 (0.223) |
| Age (years) | [11; 30] | 86 | 68 |  |  |  |
|  | [31; 88] | 95 | 93 | 1.27 | 0.81 - 1.90 | 1.08 (0.328) |
| Characteristics |  | Knowledge of mosquito larvae |  |  |  |  |
| | | Yes | No | OR | 95% CI | $\chi^2$ (p) |
| Education level | < Primary | 6 | 48 |  |  |  |
|  | Primary and more | 43 | 245 | 0.71 | 0.28 - 1.77 | 0.73 (0.464) |
| Areas | Peri-urban | 7 | 67 |  |  |  |
|  | Urban | 42 | 226 | 0.56 | 0.24 - 1.31 | 1.34 (0.182) |
| Gender | Female | 22 | 153 |  |  |  |
|  | Male | 27 | 140 | 0.75 | 0.40 - 1.37 | 0.95 (0.344) |
| Age (years) | [11; 30] | 13 | 141 |  |  |  |
|  | [31; 88] | 36 | 152 | 0.39 | 0.19 - 0.76 | 2.74 (0.006) |
| Characteristics |  | Presence of positive Aedes larval breeding sites |  |  |  |  |
| | | Yes | No | OR | 95% CI | $\chi^2$ (p) |
| Type of construction | Traditional & semi-modern | 171 | 62 |  |  |  |
|  | Modern | 159 | 122 | 2.12 | 1.45 - 3.08 | 3.93 (<0.001) |
| Knowledge of the vector |  |  |  |  |  |  |
| Areas | Peri-urban | 80 | 39 |  |  |  |
|  | Urban | 250 | 145 | 1.12 | 0.77 - 1.84 | 0.79 (0.433) |
| Annual income | < 1,200,000 FCFA | 187 | 75 |  |  |  |
|  | ≥ 1,200,000 FCFA | 143 | 109 | 0.53 | 0.36 - 0.76 | 3.44 (<0.001) |
| Water supply source | Public fountain | 102 | 49 |  |  |  |
|  | Tap water/Well | 228 | 135 | 1.23 | 0.82 - 1.85 | 1.02 (0.307) |
| Residents/concession | [11; 38] | 113 | 40 |  |  |  |
|  | [1; 10] | 217 | 144 | 1.88 | 1.23 - 2.85 | 2.95 (0.003) |

The glmm analysis displayed in Table 7 shows that knowledge of the dengue vector was better in the HD of Bogodogo than in the reference HD of Baskuy, and in Nongremasson. Knowledge of the dengue vector was also associated with respondent age, sex, and occupation. Finally, the economic level and type of construction of respondents’ compound were found to be associated with the level of general knowledge about dengue fever.

**Table 7:** Associations between the dengue vector knowledge level and different characteristics of the visited compound, respondents’ socioeconomic status and health district following a Generalised Linear Model (glm). Significant predictors are bolded.

| Predictors | Estimates | CI | p |
| --- | --- | --- | --- |
| **(Intercept)** | **5.78** | **5.16 – 6.41** | **<0.001** |
| **HD [Baskuy]** | - | - | - |
| **Bogodogo** | **0.99** | **0.80 – 1.18** | **<0.001** |
| Nongremassom | 0.13 | -0.06 – 0.33 | 0.186 |
| **Sex [F]** | - | - | - |
| **M** | **-0.32** | **-0.49 – -0.15** | **<0.001** |
| **Age** | **0.01** | **0.00 – 0.02** | **0.002** |
| **Instruction level [No education]** | - | - | - |
| No formal schooling | -0.33 | -0.79 – 0.13 | 0.158 |
| **Primary school** | **-0.32** | **-0.60 – -0.03** | **0.028** |
| Secondary | -0.21 | -0.49 – 0.07 | 0.147 |
| University | 0.27 | -0.07 – 0.62 | 0.115 |
| **Occupation [Craftsmen]** |  |  |  |
| **Other** | **-0.79** | **-1.15 – -0.44** | **<0.001** |
| **Trader** | **-0.87** | **-1.12 – -0.61** | **<0.001** |
| Government employees | -0.06 | -0.34 – 0.23 | 0.702 |
| **Housewife** | **-1.05** | **-1.35 – -0.75** | **<0.001** |
| **Student** | **-1.05** | **-1.31 – -0.78** | **<0.001** |
| **Annual income [I]** | - | - | - |
| **II** | **-0.85** | **-1.26 – -0.45** | **<0.001** |
| **III** | **-0.75** | **-1.16 – -0.33** | **<0.001** |
| **IV** | **-0.87** | **-1.29 – -0.45** | **<0.001** |
| **House type [Mixed]** | - | - | - |
| **Modern** | **0.36** | **0.16 – 0.56** | **<0.001** |
| **Traditional** | **-0.64** | **-0.90 – -0.39** | **<0.001** |

Building materials used in the construction of the compound and the number of residents were associated with the presence of potential larval containers, while the presence of immature positive containers is affected only by the number of residents (S3Table, S4Table). Traditional and semi- modern type compounds had a higher likelihood of containing positive breeding sites with immature forms of *Ae. aegypti* (OR = 2.12; 95% CI=1.45 - 3.08; p < 0.001) compared to modern type compounds (Table 5). Similarly, compounds with more than 10 residents were more likely to harbour positive breeding sites (OR = 1.88; 95% CI=1.23 - 2.85; p = 0.003) compared to those with ten or fewer residents.

### Larval breeding sites and Stegomyia indices

A wide variety of water-holding containers (potential larval sites) were identified in the compounds during our entomological survey. These containers were categorized by type based on their similarity or purpose (Table 8 and 9). The most commonly found larval sites were tires and buckets/cans/pots (BCP), followed by drums/barrels (DB) (Table 8 and 9). Out of a total of 2290 potential larval sites documented in the study area, 29.69% were tires and 28.86% were BCP. Tires accounted for between 26% and 34.25% of the potential breeding sites inn each HD, while BCP accounted for between 27% and 30.37% (Table 8). These containers were the most commonly found in both urban and peri-urban areas (Table 9). In the urban area, tires were prevalent in 31.13% of cases, compared to 24.76% in the peri-urban area. Similarly, BCP, accounted for 27.98% in the urban area and 31.91% in the peri-urban area as potential larval breeding sites.

**Table 8.**
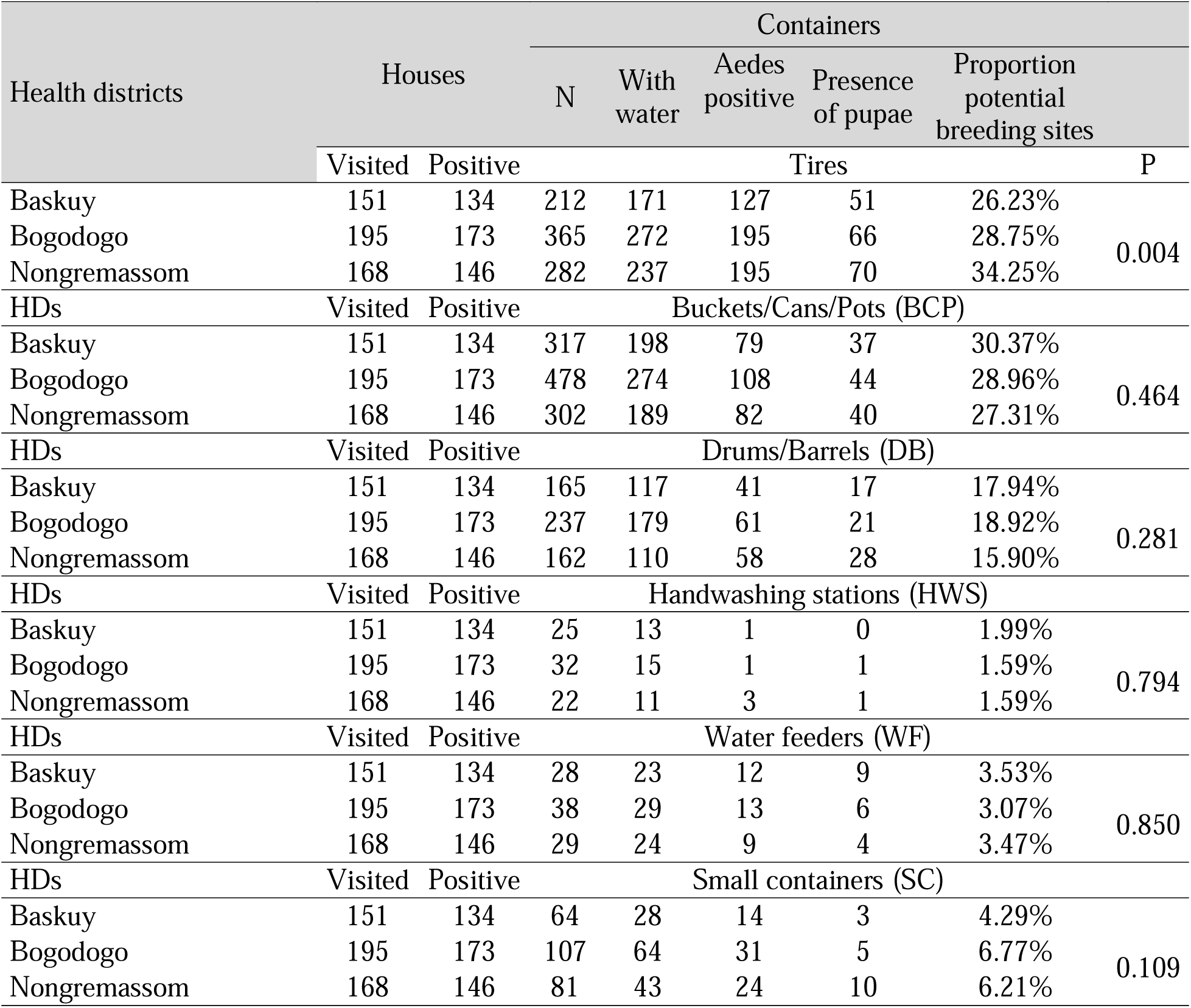
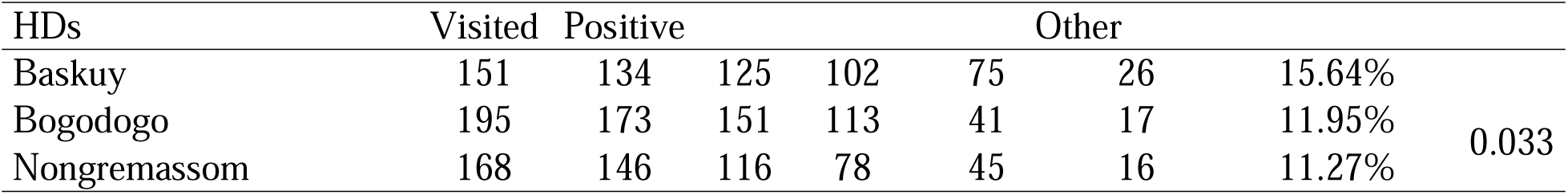
Distribution of containers, potential larval breeding sites and larval breeding sites by type and according to health district.

**Table 9.**
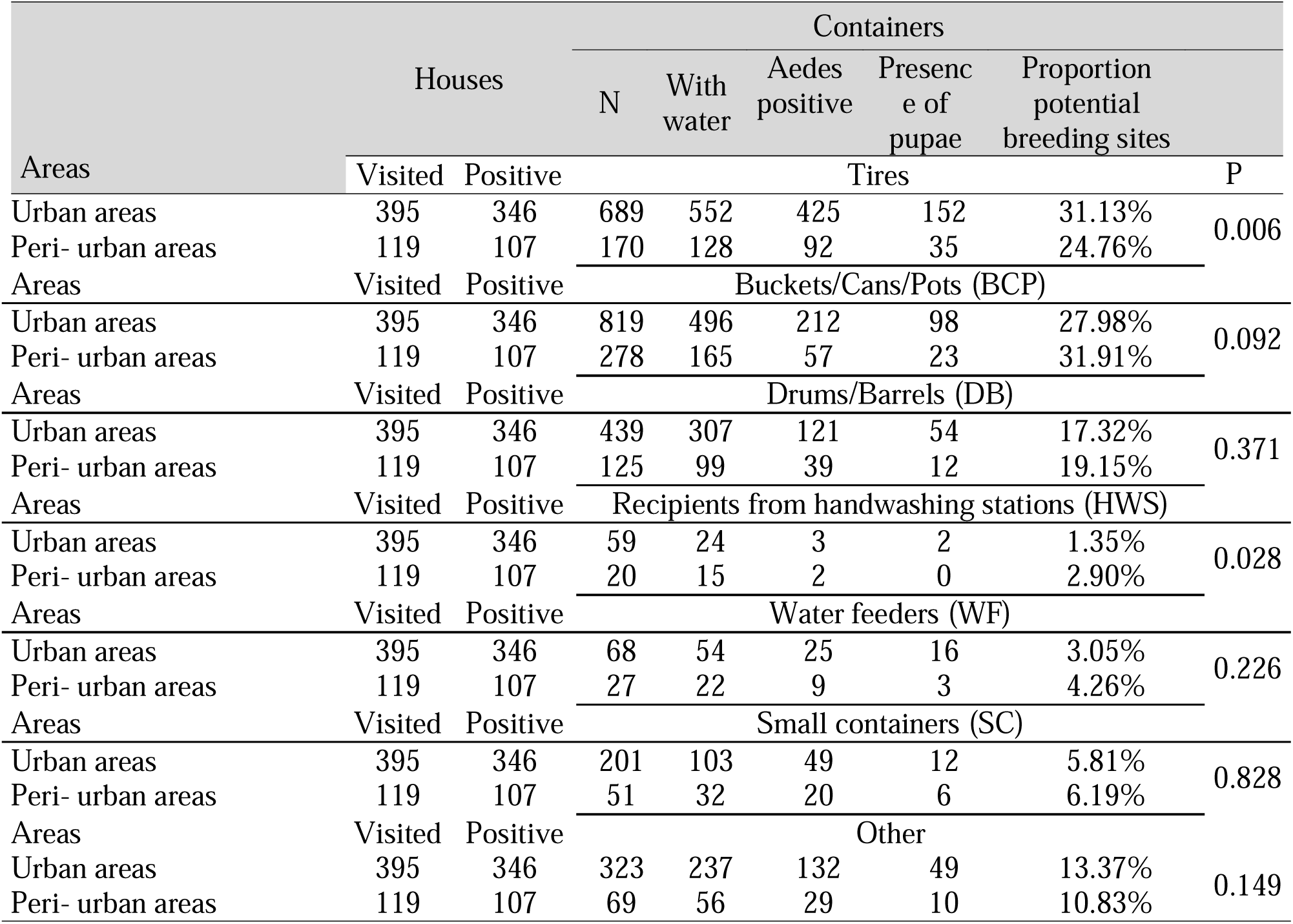
Distribution of containers, potential larval sites and larval sites by type and according to type of areas.

Stegomyia indices for the HDs and zones (urban and peri-urban) are reported in Table 8. The HDs of Baskuy, Bogodogo, and Nongremassom displayed house indices (HIs) of 88.7%, 88.7%, and 86.9% respectively, along with container indices (CI) of 53.5%, 47.6%, and 60.1%. Regarding the Breteau indices (BI), 231.1, 230.8, and 247.6 were recorded respectively in the HDs of Baskuy, Bogodogo, and Nongremassom.

In the urban and peri-urban areas, the HIs were 87.6% and 89.9%, respectively, the CIs were 54.5% and 48.0%, and the BIs were 244.8 and 208.4 (Table 10).

**Table 10:** Entomological (Stegomyia) indices according to health districts (HD) and type of areas (Urban and Peri-urban)

| HDs and areas | Houses |  | Containers |  | HI (%) | CI (%) | BI |
| --- | --- | --- | --- | --- | --- | --- | --- |
|  | Visited | Positive | With water | Aedes positive |  |  |  |
| HD Baskuy | 151 | 134 | 652 | 349 | 88.7 | 53.5 | 231.1 |
| HD Bogodogo | 195 | 173 | 946 | 450 | 88.7 | 47.6 | 230.8 |
| HD Nongremassom | 168 | 146 | 692 | 416 | 86.9 | 60.1 | 247.6 |
| Urban areas | 395 | 346 | 1773 | 967 | 87.6 | 54.5 | 244.8 |
| Peri-urban areas | 119 | 107 | 517 | 248 | 89.9 | 48.0 | 208.4 |
| Total | 514 | 453 | 2290 | 1215 | 88.1 | 53.1 | 236.4 |

The recorded HIs showed no significant difference between HDs (p = 0.835) and between the urban and peri-urban areas (p = 0.599). However, CIs were significantly different not only between HDs (p < 0.001) but also between urban and peri-urban areas (p = 0.009).

### Risk factors for dengue vector proliferation

Based on the glmm analyses (S2Table, S3Table, and S4Table), the occurrences of Aedes potential breeding containers, *Aedes* immature-positive breeding containers, and *Aedes* pupae-positive containers are associated with various factors. Compounds with type II, III, and IV levels of annual income were negatively correlated with the presence of *Aedes* pupae-positive containers (S2Table), *Aedes* larvae-positive containers (S3Table), and *Aedes* potential breeding sites containers compared to the reference type I. The presence of *Aedes* potential breeding containers and *Aedes* immature- positive breeding sites containers is linked to the number of residents per compound (S3Table and S4Table). Our findings suggest a connection between the population’s knowledge level of dengue vectors and the presence of potential larval containers and pupae-positive containers (S2Table and S3Table). However, this association is slightly positive with potential breeding site containers and negative with pupae-positive containers.

Finally, our GLMM analysis demonstrates a positive association between the presence of pupae- positive breeding sites in the HD of Nongremassom compared to the reference, Baskuy (S2Table).

## Discussion

Since 2013, Burkina Faso has experienced outbreaks of dengue cases, especially in the city of Ouagadougou where the main vector of this arbovirus, *Ae. aegypti*, is well established [29,30]. This socio-entomological study has helped identifying the level of knowledge and practices of the population regarding dengue and its vector in the health districts of Baskuy, Bogodogo, and Nongremassom. It also highlighted the level of dengue vector infestation in the compounds, as well as the types of larval containers and associated factors. The Stegomyia indices were also recorded for all HDs.

### Respondents’ knowledge of dengue

A significant proportion (94.36%) of the surveyed population in the HDs reported having heard of dengue, echoing results from similar studies conducted in endemic regions. Studies conducted in Delhi [31] and Dhaka, Bangladesh [16,32] also revealed that over 90% of respondents were familiar with dengue. Quintero *et al*. [33] noted in a multi-country study in Latin America that more than 97% of participants had knowledge of dengue and a recent study in Bangladesh by Hossain *et al*. [34] reported a similar awareness rate of 93.8%, comparable with Burkina Faso where dengue emergence is recent.

The recent outbreaks in Burkina Faso have likely contributed to spreading information about dengue among our study population [35,36]. Increased awareness about the disease would explain why respondents reported having heard about dengue through radio (42.27%), television (46.39%), relatives and friends (53.40%), and at health centres (27.42%). These information channels were also recognized as major information source in other studies [34,37]. Moreover, our study revealed significant associations between the knowledge level of dengue and characteristics such as gender and age. Female respondents and those younger than 30 years were the most likely to be unaware of dengue. This aligns with the findings of Selvarajoo *et al*. [38]who also observed that dengue knowledge was associated with age.

Although the respondents had heard of dengue fever, many were still unaware of certain aspects of this infection, such as the symptoms and mode of transmission. Fever and headaches were known by roughly half of the respondents as dengue symptoms. In many similar studies, response rates towards these two major symptoms (fever and headache) are very high [37,39]. The odds ratios revealed a positive association between the level of education and knowledge of dengue symptoms and mode of transmission. Our results align with other studies showing that knowledge about dengue transmission and symptoms is linked to the respondent’s education level [15,39]. However, Kumaran et al. [39] found that, unlike our results, female respondents were more likely to have knowledge about dengue symptoms.

A thorough understanding of dengue transmission among the population is essential for disease prevention and control efforts. Approximately 70% of the respondents were aware that dengue is transmitted by mosquitoes. However, a study on population vulnerability to dengue in Ouagadougou revealed that only 62% of respondents knew about dengue transmission by a mosquito [21]. This discrepancy could be attributed to the timing of the study in Ouagadougou, conducted shortly after the first dengue outbreak, indicating lower knowledge levels compared to our study in 2021. Recent studies in endemic regions of South America and Asia have shown that a significant percentage of respondents (90 - 100%) were knowledgeable about dengue transmission by mosquito vectors [34,38]. Disparities in the effectiveness of health and public hygiene systems among countries, responsible for information dissemination, education, and communication activities, along with health education strategies, are likely to influence the population’s understanding of diseases and their modes of transmission [40,41].

### Respondents’ knowledge of the dengue vector

Only 36.55% (125/342) of the respondents aware of dengue transmission modes knew of at least one breeding site of the vector. This low rate poses a significant obstacle to dengue control. Studies conducted in Cambodia and Yemen have reported a higher level of knowledge of breeding sites, with rates ranging from 86.1 to 95.5% [39,42].

Knowledge of a vector’s host-seeking periods is essential for bite prevention. Our study revealed that 75.73% of respondents had knowledge of the vector host-seeking time. This finding is consistent with previous reports by Udayanga et al. [20] in Colombo, Sri Lanka but lower than the level found by Rahman et al. [43,44] in Thailand, country that has experienced multiple dengue outbreaks since the 1950s [45]

If knowledge of the adult dengue vector may have implications for mosquito bite control or prevention, understanding the larval stages in breeding sites would be even more crucial. The lack of knowledge of the mosquito larval stage is likely to contribute to the proliferation of breeding sites and, consequently, adult mosquitoes. Our results revealed that only 32.16% and 14.16% of the respondents could recognize the adult mosquito of the dengue vector and mosquito larvae, respectively. In Brazil and Colombia, a study by Laurentino [46] reported a significantly higher (88%) level of knowledge about dengue vector compared to our study which was based on visual recognition of the mosquitoes. Similarly, a multi-country study in urban and peri-urban Asia (India, Indonesia, Myanmar, Philippines, Sri Lanka, and Thailand) on the eco-bio-social determinants of dengue vector reproduction, found a much higher level of knowledge about mosquito larvae (47% to 97%) than our study [17], that may again be related to the methodology based on survey, compared to visual recognition of mosquito larvae.

### Practices that may affect dengue transmission

Some of the population’s practices may favour vector proliferation and disease transmission. For example, over 50% of sampled compounds have water storage containers, particularly in peri-urban areas where this is observed in 80% of the compounds. This is influenced by the water supply system in cities [47,48], more prevalent in peri-urban areas with scarce tap water: year-round water storage in compounds poses a risk for dengue vector proliferation and potential disease transmission [49]. However, many individuals are unaware that water storage containers can serve as a breeding site for the dengue vector [16].

In addition, 73.35% of respondents reported not habitually emptying containers with unused water. This suggests that few respondents adhere to proper practices by emptying non-useful water containers, which can serve as larval sites for *Aedes* mosquitoes, particularly in the rainy season. This percentage is lower compared to results from other studies and contributes to the spread of the dengue vector [42,50]

### Knowledge of mosquito bite prevention

Knowledge and practices of mosquito bite prevention are crucial for individuals to protect themselves from vector-borne diseases such as malaria and dengue. Our study revealed that two-thirds of the sampled compounds lacked mosquito screens on their windows. Although the effectiveness of installing window screens in preventing mosquito bites has been proven [27,50], it is important to note that, this effectiveness is closely related to contextual mosquito behaviour and may not have a significant impact on the exophilic and exophagous mosquito population of Ouagadougou [12].

As for protection against mosquito bites, over 80% of our respondents reported sleeping under a bed net at night, while only about 17% did so during the day. In a KAP study on the dengue vector in Thailand, Koenraadt *et al.* [15] reported a similar proportion (18%) of respondents slept under bed nets during the day. The use of other prevention strategies against mosquito bites, such as mosquito repellents, lotions, fog, spray cans, fans, long-sleeved clothing, destroying breeding sites, emptying water containers, covering water storage containers, and others, was reported in lower proportions but has been reported in various studies [20,34].

### *Aedes aegypti* mosquito larval sites

Significant domestic infestation, with a wide variety of potential larval breeding containers, was also observed. The types of potential larval containers in both the HDs and the urban and peri-urban areas were found to be similar. The primary breeding sites identified were used tires, followed by BCP and water storage containers [11,51]. The increasing global use of cars has led to a rise in tire trade between countries and continents [21]. When these used tires are exposed to rain, they become potential larval sites for *Aedes* breeding due to the conducive environment they provide [52]

Larval containers contribute not only to increasing the population density of vector mosquitoes but also to the risk of arbovirus transmission. Our results, as indicated, show a negative association between the type of construction and the presence of positive breeding sites, while a positive association was found with the number of people living in the compound surveyed, a phenomenon also documented by Walker et al. [53]. The greater the number of residents, the more waste would be produced, some of which may act as rainwater containers and become potential larval containers in the compound. Furthermore, our findings reveal that households with higher income levels were less likely to host potential mosquito larval containers and even less likely to have positive containers for larvae or pupae. These results align with those of Walker et al. [53], who also demonstrated a negative association between compound income and infestation with mosquito larval containers.

The Stegomyia indices calculated in our study were notably higher compared to those in other studies [53,54]. The purpose of water storage in this context is not solely for immediate use but rather to supplement unpredictable water shortages. Across all three HDs in the study area, the Stegomyia indices, including HI, CI, and BI (Tables 9), exceeded the threshold values recognized by the Pan American Health Organization and WHO [55,56]. These values strongly indicate the population’s limited knowledge about recognizing particularly Aedes larval sites and recognising mosquito larvae, and the associated diseases, pointing towards a heightened risk of arbovirus transmission when the pathogen is present in the study area [28].

## Conclusion

In spite of the multiple sensitizations through messages on television, radio and news journals on dengue and its transmission, following the outbreaks that the country has experienced, our findings showed that knowledge about certain aspects of dengue, especially its vector is still partial. Our findings also indicate practices and attitudes that could favour the proliferation of dengue vectors. In addition, these findings highlight a high number of potential larval containers of various types, and very high Stegomyia indices in the study area, suggesting a significant infestation of the compounds by the dengue vector. It is essential to increase communication and awareness activities on dengue through campaigns or other channels aimed at improving practices and attitudes towards prevention and also to enhance the level of knowledge of the population on the disease and its transmission. This could ultimately enhance the community’s knowledge and awareness and help better involve the population in the control of dengue and other mosquito-borne diseases in general.

## Data Availability

All data produced in the present study are available upon reasonable request to the authors

---

**Supplementary Table 1:**
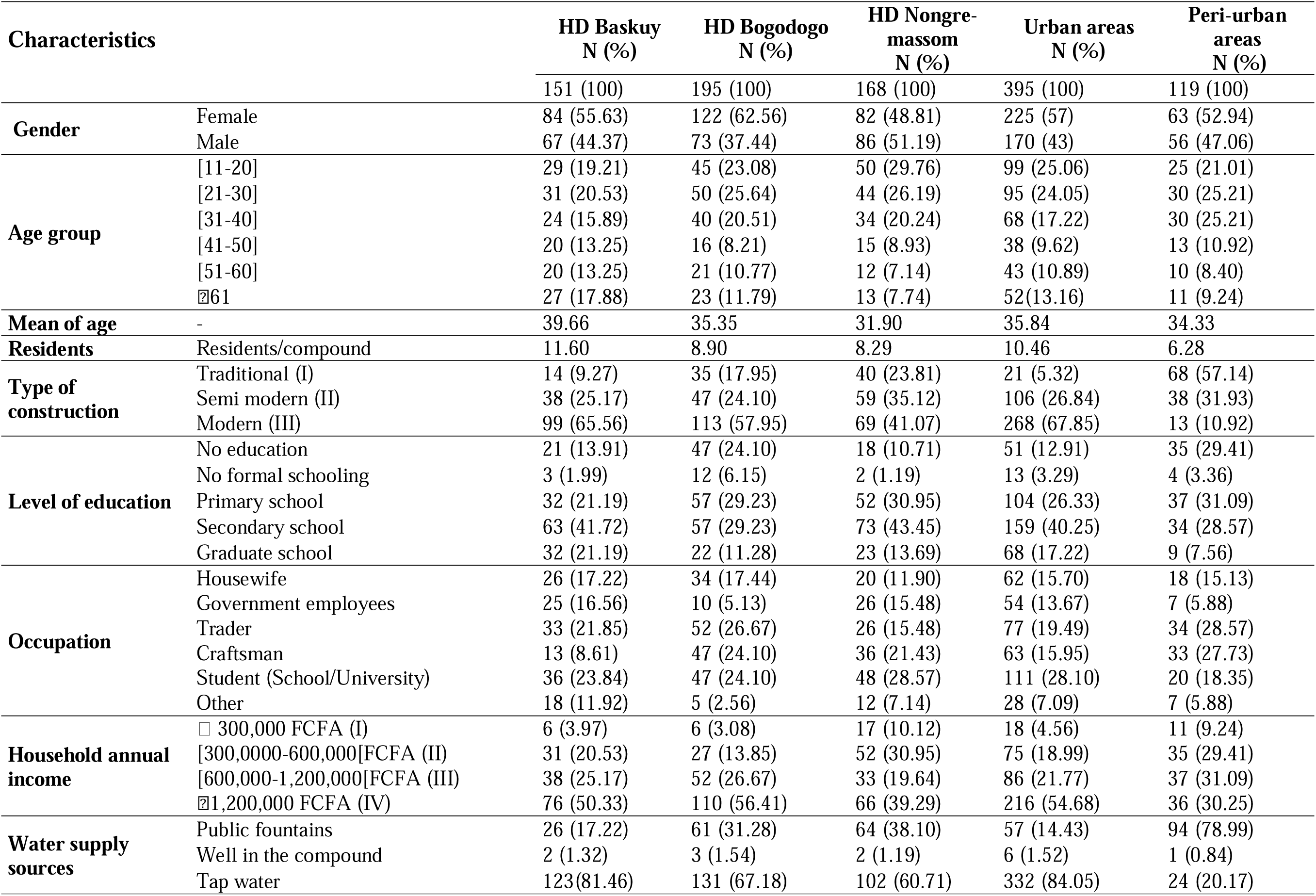
Socio-demographic and economic characteristics of the respondents and compounds surveyed.

**Supplementary Table 2:** Table showing the associations between the presence of pupae positive breeding sites containers and different characteristics of the visited compound, its location and respondents knowledge level following Generalized Linear Mixed Model analysis (GLMM).

| Predictors | Incidence Rate Ratios | CI | p |
| --- | --- | --- | --- |
| (Intercept) | 0.29 | 0.14 – 0.61 | **0.001** |
| HD [Baskuy] | - | - | - |
| Bogodogo | 0.83 | 0.55 – 1.24 | 0.356 |
| **Nongremassom** | **1.53** | **1.06 – 2.22** | **0.024** |
| Annual income [I] | - | - | - |
| **II** | **0.46** | **0.25 – 0.84** | **0.013** |
| **III** | **0.42** | **0.23 – 0.79** | **0.007** |
| **IV** | **0.28** | **0.16 – 0.51** | **<0.001** |
| Residents per house | 1.02 | 1.00 – 1.04 | 0.103 |
| **Population knowledge on dengue vector** | 0.92 | 0.85 – 1.00 | **0.047** |

**Supplementary Table 3:**
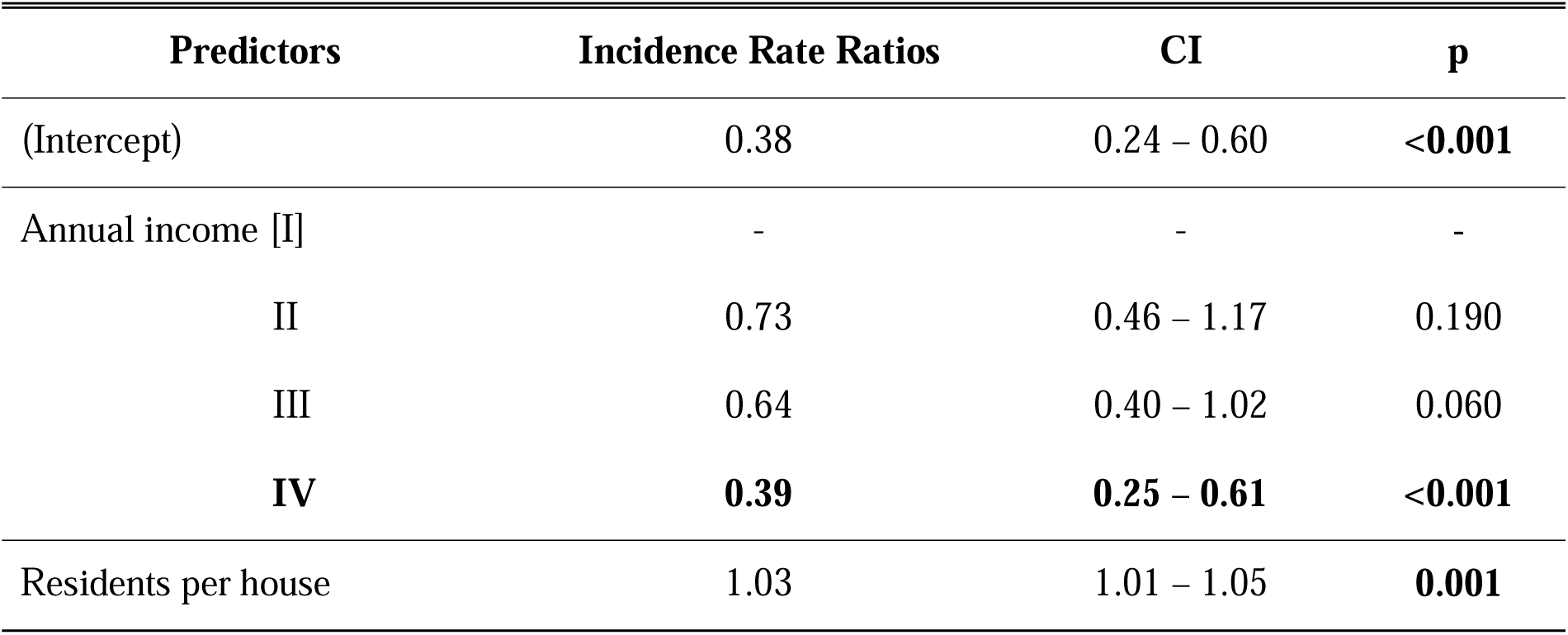
Table showing the associations between the presence of positive Aedes mosquito immature breeding sites containers and different characteristics of the visited compound and respondents knowledge level following Generalized Linear Mixed Model analysis (GLMM).

**Supplementary Table 4:** Table showing the associations between the presence of potential larval containers and different characteristics of the visited compound and respondents knowledge level following the Generalized Linear Mixed Model analysis (GLMM).

| Predictors | Incidence Rate Ratios | CI | p |
| --- | --- | --- | --- |
| **(Intercept)** | **0.61** | **0.37 – 0.99** | **0.047** |
| Annual income [I] | - | - | - |
| **II** | **0.64** | **0.42 – 0.97** | **0.036** |
| III | 0.76 | 0.50 – 1.16 | 0.200 |
| **IV** | **0.65** | **0.42 – 1.00** | **0.050** |
| House type [Semi-traditional] | - | - | - |
| **Modern** | **0.72** | **0.58 – 0.89** | **0.002** |
| Traditional | 1.02 | 0.77 – 1.34 | 0.903 |
| **Residents per house** | **1.02** | **1.01 – 1.03** | **0.003** |
| **Population knowledge on dengue vector** | **1.07** | **1.02 – 1.11** | **0.003** |

